# Routine laboratory blood tests predict SARS-CoV-2 infection using machine learning

**DOI:** 10.1101/2020.06.17.20133892

**Authors:** He S. Yang, Yu Hou, Ljiljana V. Vasovic, Peter Steel, Amy Chadburn, Sabrina E. Racine-Brzostek, Priya Velu, Melissa M. Cushing, Massimo Loda, Rainu Kaushal, Zhen Zhao, Fei Wang

## Abstract

**Background:** Accurate diagnostic strategies to rapidly identify SARS-CoV-2 positive individuals for management of patient care and protection of health care personnel are urgently needed. The predominant diagnostic test is viral RNA detection by RT-PCR from nasopharyngeal swabs specimens, however the results are not promptly obtainable in all patient care locations. Routine laboratory testing, in contrast, is readily available with a turn-around time (TAT) usually within 1-2 hours.

**Method:** We developed a machine learning model incorporating patient demographic features (age, sex, race) with 27 routine laboratory tests to predict an individual’s SARS-CoV-2 infection status. Laboratory test results obtained within two days before the release of SARS-CoV-2-RT-PCR result were used to train a gradient boosted decision tree (GBDT) model from 3,356 SARS-CoV-2 RT-PCR tested patients (1,402 positive and 1,954 negative) evaluated at a metropolitan hospital.

**Results:** The model achieved an area under the receiver operating characteristic curve (AUC) of 0.854 (95% CI: 0.829-0.878). Application of this model to an independent patient dataset from a separate hospital resulted in a comparable AUC (0.838), validating the generalization of its use. Moreover, our model predicted initial SARS-CoV-2 RT-PCR positivity in 66% individuals whose RT-PCR result changed from negative to positive within two days.

**Conclusion:** This model employing routine laboratory test results offers opportunities for early and rapid identification of high-risk SARS-CoV-2 infected patients before their RT-PCR results are available. It may play an important role in assisting the identification of SARS-COV-2 infected patients in areas where RT-PCR testing is not accessible due to financial or supply constraints.

## Introduction

The Coronavirus Disease-2019 (COVID-19) pandemic has rapidly spread worldwide resulting in over 14 million confirmed cases and more than 603,000 total deaths as of July 20, 2020 *(1)*. The highly contagious nature of SARS-CoV-2 *(2)*, rapid progression of disease in some infected patients *(3)* and the subsequent stress on the healthcare system has created an urgent need for rapid and effective diagnostic strategies for the prompt identification and isolation of infected patients. Currently, the diagnosis of COVID-19 relies on SARS-CoV-2 virus-specific real-time reverse-transcriptase polymerase chain reaction (RT-PCR) testing of nasopharyngeal swabs or other upper respiratory track specimens *(4, 5)*. However, while the TAT of RT-PCR testing is usually within 48 hours *(6)*, it can be substantially longer due to many variables including the need for repeat testing or the lack of needed supplies. Many smaller hospitals do not yet have access to on-site SARS-CoV-2 RT-PCR testing. These issues can result in delayed hospital admission and bed assignment, inappropriate medical management including quarantining of infected patients and increased exposure of healthcare personnel and other patient contacts to the virus. Rapid diagnosis and identification of high-risk patients for early intervention is vital for individual patient care, and, from a public health perspective, for controlling disease transmission and maintaining the healthcare workforce.

Currently in hospital EDs, the nationally recommended practice when evaluating patients with moderate to high risk for COVID-19 is SARS-CoV-2 RT-PCR testing, a panel of routine laboratory tests, a chest X-ray and symptomatology, whereas chest computed tomography (CT) is not recommended due to cost and TAT considerations *(2, 7)*. Routine laboratory tests are generally available within 1-2 hours and are accessible prior to patient discharge from the ED. Several studies *(3, 8-10)* have reported laboratory abnormalities in COVID-19 patients on admission and during the disease course, including increases in C-reactive protein (CRP), D-dimer, lactic acid dehydrogenase (LDH), cardiac troponin, procalcitonin (PCT), and creatinine as well as lymphopenia and thrombocytopenia. While no single laboratory test can accurately discriminate SARS-CoV-2 infected from non-infected patients, the combination of the results of these routine laboratory tests may predict the COVID-19 infection status.

Recent promising advances in the application of artificial intelligence (AI) in several healthcare areas *(11-15)* have inspired the development of AI-based algorithms as diagnostic *(6)* or prognosis tools *(16)* for complex diseases, such as COVID-19. In this study, we hypothesized that the results of routine laboratory tests performed within a short time frame as the RT-PCR testing, in conjunction with a limited number of previously identified predictive demographic factors (age, gender, race) *(17)*, can predict SARS-CoV-2 infection status. Thus, we aimed to develop a machine learning model integrating age, gender, race and routine laboratory blood tests, which are readily available with a short TAT.

## Methods

### Data collection

We conducted a retrospective study with 5,893 patients evaluated at the New York Presbyterian Hospital/Weill Cornell Medicine (NYPH/WCM) during March 11 to April 29, 2020. SARS-CoV-2 RT-PCR results, routine laboratory testing results and patient demographic information were obtained from the laboratory information system (Cerner Millennium, Cerner Corporation). Exclusion criteria included patients < 18 years old, patients who had indeterminate RT-PCR results, and patients who did not have laboratory results within two days prior to the completion of RT-PCR testing (**Figure 1**). Among a total of 4,207 RT-PCR results from 1,402 RT-PCR positive and 1,954 negative patients in our dataset, 54.1% of RT-PCR tests were ordered from the Emergency Department (ED), 32.4% were ordered on inpatients, including 2.7% from ICU patients, and the rest were ordered from the outpatient surgery department, the outpatient clinics, and the private ambulatory setting. Among the RT-PCR results excluded from the dataset due to no corresponding laboratory results, 50.0% were ordered for “non-patient institutional” including patient samples used for validation, plasma donor specimens, and healthcare workers. An additional 20.0% of excluded RT-PCR tests were ordered on ED patients who were likely discharged, 12% were from “private ambulatory” setting, 2.8% were ordered for outpatient clinic with no associated hospital admission, and the rest were from the surgery or dental departments to rule out COVID-19 infection. Same criteria were applied to the dataset collected from New York Presbyterian Hospital/Lower Manhattan Hospital (NYPH/LMH) during the same time period. A total of 1,822 RT-PCR tests ordered for 496 RT-PCR positive and 968 negative NYPH/LMH patients were obtained and used for validation. Among them, 60.9% were ordered from ED and 36.3% were from inpatients. This study was approved by the Institutional Review Board (#20-03021671) of Weill Cornell Medicine.

**Figure 1.**
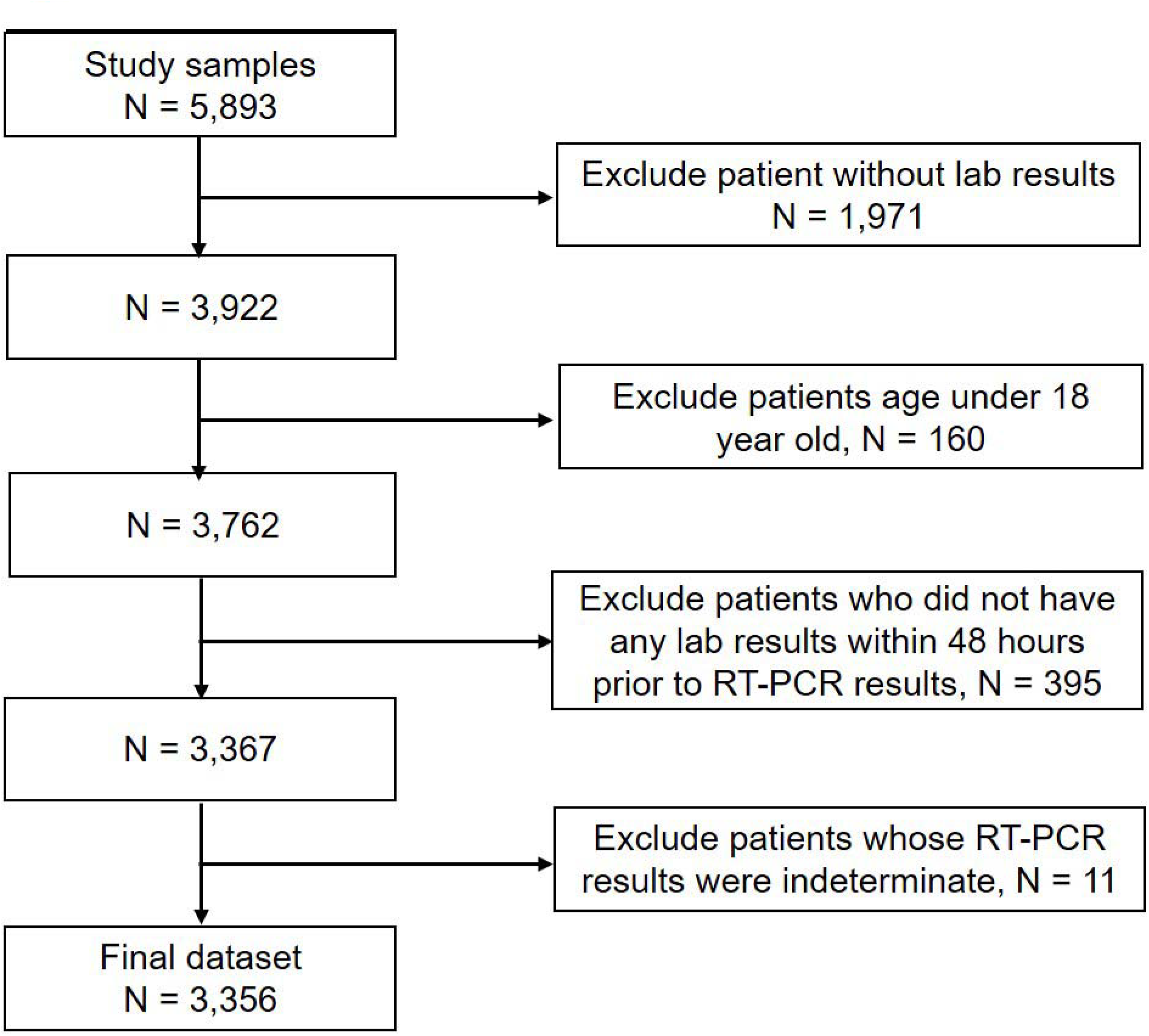
Inclusion/exclusion cascade of patients in the dataset.

### SARS-CoV-2 RT-PCR testing

SARS-CoV-2 RT-PCR testing was performed at NYPH/WCM using the RealStar SARS-CoV-2 RT-PCR kit 1.0 (Altona Diagnostics) reagent system which targets on the S gene and E gene, the Cobas SARS-CoV-2 RT-PCR assay (Roche Diagnostics) which targets on the ORF1 a/b gene and E gene, the Panther Fusion SARS-CoV-2 RT-PCR assay (Hologic) which targets on the OFR1 a/b gene region 1 and region2, and Xpert Xpress SARS-CoV-2 RT-PCR (Cepheid) which targets on the N2 gene and E gene *(18)*. Among 4,207 RT-PCR reactions included in the NYPH/WCM training set, 1187 (28.2%), 1768 (42.0%), 3 (0.0%), and 1249 (29.7%) were performed on the Altona, Roche Cobas, Hologic, and Cepheid platform, respectively. Based on the laboratory validation, there was no meaningful difference in the sensitivity of each platform *(19, 20)*. There was no difference in turnaround time for positive or negative RT-PCR results. RT-PCR was performed using the Roche Cobas SARS-CoV-2 RT-PCR assay and Cepheid Xpert Xpress SARS-CoV-2 RT-PCR at NYPH/LMH.

### Routine Laboratory testing

At NYPH/WCM, routine chemistry testing was performed on Siemens ADVIA XPT analyzers and Centaur XP analyzers. Procalcitonin was performed on the Roche e411 analyzer. Blood gas analysis was performed on the Instrumentation Laboratory GEM Premier 4000 analyzer. Routine hematological testing was performed on the UniCel DXH 800 analyzer. Coagulation tests were performed on the Instrumentation Laboratory ACLTM TOP CTS Coagulation System.

At NYPH/LMH, Routine chemistry testing including procalcitonin was performed on Abbott ARCHITECT® c SYSTEM ci 4100 and ci 8200 analyzers. Blood gas analysis was performed on the Radiometer analyzer ABL 820 FLEX. Routine complete blood count (CBC) testing was performed on the UniCel DXH 800 analyzer. Coagulation tests were performed on the STAGO STA-R® Evolution multiparametric analyzer.

### Model construction

A total of 685 distinct laboratory tests were ordered for patients in the NYPH/WCM dataset. A 685-dimensional vector was generated for each RT-PCR test. If one specific test was ordered multiple times, an average of the values was calculated and used for analysis. Univariate analysis was performed on all laboratory test results to obtain the significance of the association between each laboratory test and the RT-PCR result with SciPy1.4.1 *(21)*. Laboratory tests were selected to construct the input feature vectors of the prediction model based on the following criteria: 1) a result available for at least 30% of the patients two days before a specific SARS-CoV-2 RT-PCR test, and 2) showing a significant difference (*P*-value, *P*-value after Bonferroni correction, *P*-value after demographics adjustment all less than 0.05) between patients with positive and negative RT-PCR results. After the feature selection process (details are provided in the online **Supplemental Material**), a 33-dimensional vector (27 routine lab tests, one age, one gender, four race variables (African American, Asian, Caucasian and others) was constructed to represent every RT-PCR test. The value on each dimension was the average result value of the corresponding laboratory test taken two days before the RT-PCR test in addition to the patients’ age, gender and race. The patient’s race and gender variables were encoded with binary values. The missing value of a specific laboratory test in a feature vector was imputed by the median value of the available non-missing value of that dimension over all patients. The result of each RT-PCR test was referred to as the label of the test.

Mathematically, let X_i_ be the 33-dimensional feature vector of the *i*-th RT-PCR test. Let *y_i_* ∊ {0,1} be its corresponding label. *y_i_* = 0 means the result of the *i*-th RT-PCR test is “Not Detected” and we refer to this RT-PCR test as a negative sample, while *y_i_* = 1 means the result is “Detected” and we refer to this RT-PCR test as a positive sample. Our goal was to “learn” a classification function *f* that can accurately map each *x_i_* to its corresponding *y_i_*. We considered 4 popular classifiers in this study:

- Logistic regression, where *f* is a linear function and our implementation is based on the scikit-learn package 0.23.1 using sklearn.linear_model.LogisticRegression with all default parameter settings
- Decision tree, where *f* is a classification tree and our implementation is based on the scikit-learn package 0.23.1 *(22)* with sklearn.tree.DecisionTreeClassifier with all default parameter settings. According to the document (https://scikitlearn.org/stable/modules/tree.html#tree-algorithms-id3-c4-5-c5-0-and-cart), the decision tree algorithm implemented in scikit-learn is an optimized Classification and Regression Tree (CART) algorithm *(23)*.
- Random forest, where *f* is a random forest *(24)* and our implementation is based on scikit-learn package 0.23.1 with sklearn.ensemble.RandomForestClassifier with all default parameter settings and the number of trees equal 100.
- Gradient boosted decision tree (GBDT), where *f* is a gradient boosting machine with decision tree as base learners *(25)*. Our implementation is based on scikit-learn package 0.23.1 with sklearn.ensemble.GradientBoostingClassifier with all default parameter settings and the number of trees equal 100.

The models were evaluated in two different settings. The first setting was a 5-fold cross validation with the NYPH/WCM data, where all RT-PCR tests were randomly partitioned into 5 equal buckets with the same positive/negative ratio in each bucket as the ratio over all tests. The implementation was based on scikit-learn package 0.23.1*(22)* with the sklearn.model_selection.StratifiedKFold function. Then the training and testing procedure was performed 5 times for these 4 different classifiers. Each time a specific bucket was used for testing and the remaining 4 buckets for training. In the second setting all data from NYPH/WCM were used for training, and the data from NYPH/LMH was used for testing. In both settings, highly suspicious negatives (HSN) were excluded in the training process. Here an HSN was defined as a negative RT-PCR test in a patient who had a positive RT-PCR result upon re-testing within 2 days.

## Results

The pipeline of our modeling framework is illustrated in **Figure 2**. A summary of statistics of the 27 routine laboratory tests used to construct the input feature vectors of the prediction model is shown in online **Supplemental Table 1**. The models were trained and tested on a retrospective dataset collected from 3,356 SARS-CoV-2 RT-PCR tested adult patients who had routine laboratory testing performed within 48 hours prior to the release of RT-PCR result, between March 11 to April 29, 2020, at NYPH/WCM. This dataset included 1,402 SARS-CoV-2 RT-PCR positive and 1,954 negative patients who ranged in age from 18 to 101 years (mean 56.4 years, demographic information in **Table 1**). Among 590 patients who had repeat testing during this 7-week study period, 53 were initially negative but became positive upon repeat testing. Among this subgroup, 32 patients’ RT-PCR results changed from negative to positive within a 2-day period.

**Figure 2.**
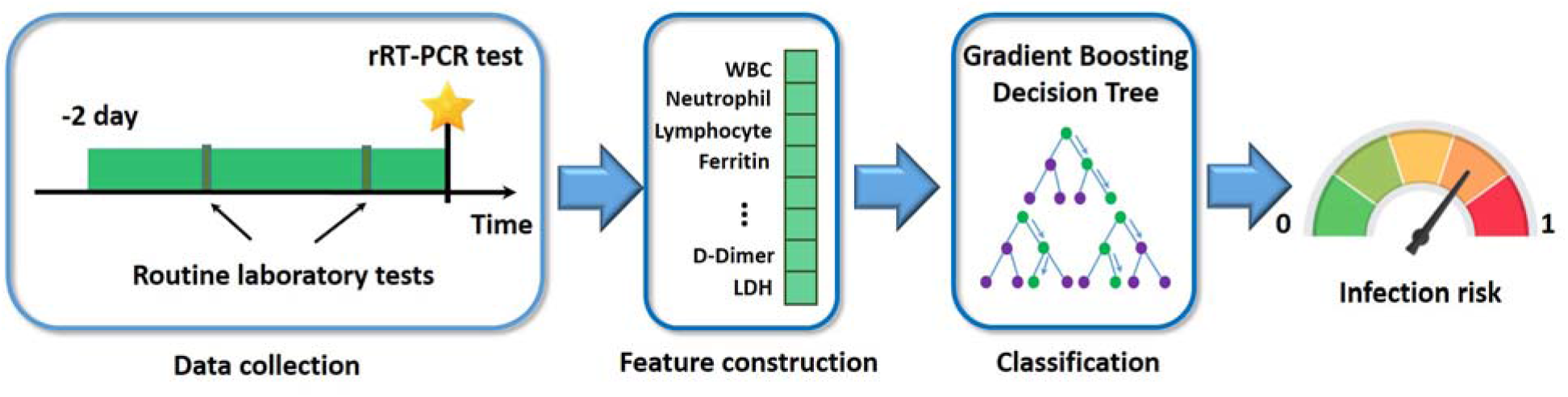
Illustration of the modeling pipeline. Routine laboratory testing results completed within two days prior to the release of RT-PCR results were used to construct a vector, upon which we built a classifier to predict the RT-PCR positive or negative result. Each dimension of the vector corresponds to a specific laboratory test, and its value corresponds to the average of all results of this laboratory test taken during the collection window. The model outputs a probability score ranging from 0 – 1, indicating the risk of SARS-CoV-2 infection.

**Table 1.** Basic demographics of the patients based on their first RT-PCR testing result.

|  | New York Presbyterian Hospital/Weill Cornell Medicine |  |  |  | New York Presbyterian Hospital/Lower Manhattan Hospital |  |  |  |
| --- | --- | --- | --- | --- | --- | --- | --- | --- |
|  | Total<br>(n = 3,356) | Positive<br>(n = 1,402) | Negative<br>(n = 1,954) | P-value<br>(Positive vs.<br>Negative) | Total<br>(n = 1,464) | Positive<br>(n = 496) | Negative<br>(n = 968) | P-value<br>(Positive<br>vs.<br>Negative) |
| **Gender** | - | - | - | 3.16e-27 | - | - | - | 2.26e-13 |
| Male | 1,558 | 805 | 753 | - | 634 | 281 | 353 | - |
| n (%) | (46.42 %) | (51.67%) | (48.33%) |  | (43.31%) | (44.32%) | (55.68%) |  |
| Female | 1,798 | 597 | 1,201 | - | 830 | 215 | 615 | - |
| n (%) | (53.58%) | (33.2%) | (66.8%) |  | (56.69%) | (25.90%) | (74.10%) |  |
| **Age** | 56.44 | 61.40 | 52.89 | 1.41e-36 | 56.20 | 65.65 | 51.36 | 2.31e-37 |
| Years (SD) | (19.46) | (17.56) | (19.98) |  | (20.81) | (18.27) | (20.37) |  |
| 19-40 years | 901 | 190 | 711 | - | 479 | 59 | 420 | - |
| n (%) | (26.85%) | (21.09%) | (78.91%) |  | (32.72%) | (12.32%) | (87.68%) |  |
| 40-60 years | 888 | 405 | 483 | - | 330 | 111 | 219 | - |
| n (%) | (26.46%) | (45.61%) | (54.39%) |  | (22.54%) | (33.64%) | (66.36%) |  |
| >60 years | 1567 | 807 | 760 | - | 655 | 326 | 329 | - |
| n (%) | (46.69%) | (51.50%) | (48.50%) |  | (44.74%) | (49.77%) | (50.23%) |  |
| **Race** | - | - | - | 5.64e-05<br>(white vs.<br>black) | - | - | - | 0.4128<br>(white vs.<br>black) |
| African American | 323 | 138 | 185 | - | 161 | 43 | 118 | - |
|  | (9.62%) | (42.72%) | (57.28%) |  | (11.00%) | (26.71%) | (73.29%) |  |
| Asian | 227 | 62 | 165 | - | 235 | 75 | 160 | - |
|  | (6.76%) | (27.31%) | (72.69%) |  | (16.05%) | (31.91%) | (68.09%) |  |
| Caucasian | 1137 | 347 | 790 | - | 259 | 59 | 200 | - |
|  | (33.88%) | (30.52%) | (69.48%) |  | (17.69%) | (22.78%) | (77.22%) |  |
| Other | 421 | 221 | 200 | - | 145 | 49 | 96 | - |
|  | (12.54%) | (52.49%) | (47.51%) |  | (9.90%) | (33.79%) | (66.21%) |  |
| Unknown | 1228 | 624 | 604 | - | 658 | 267 | 391 | - |
|  | (36.59%) | (50.81%) | (49.19%) |  | (44.95%) | (59.42%) | (41.28%) |  |

The performance of four machine learning models from 5-fold cross validation are summarized in **Figure 3**. The GBDT model achieved the best performance with an area under the receiver operating characteristic curve (AUC) value of 0.854 (95% CI: 0.829 - 0.878), sensitivity of 0.761 (95% CI: 0.744 - 0.778), specificity of 0.808 (95% CI: 0.795 – 0.821), and agreement with RT-PCR of 0.791 (0.776-0.805) at the operating point determined by the maximum Youden Index *(26)*, confidence intervals for sensitivity and specificity are analyzed by the approach based on Box-Cox transformation *(27)*. Furthermore, since 54% of RT-PCR tests were ordered from the ED, performance of the GBDT model was tested on ED patients and achieved an AUC of 0.879, a sensitivity of 0.800, a specificity of 0.825, and an agreement with RT-PCR of 0.815.

**Figure 3.**
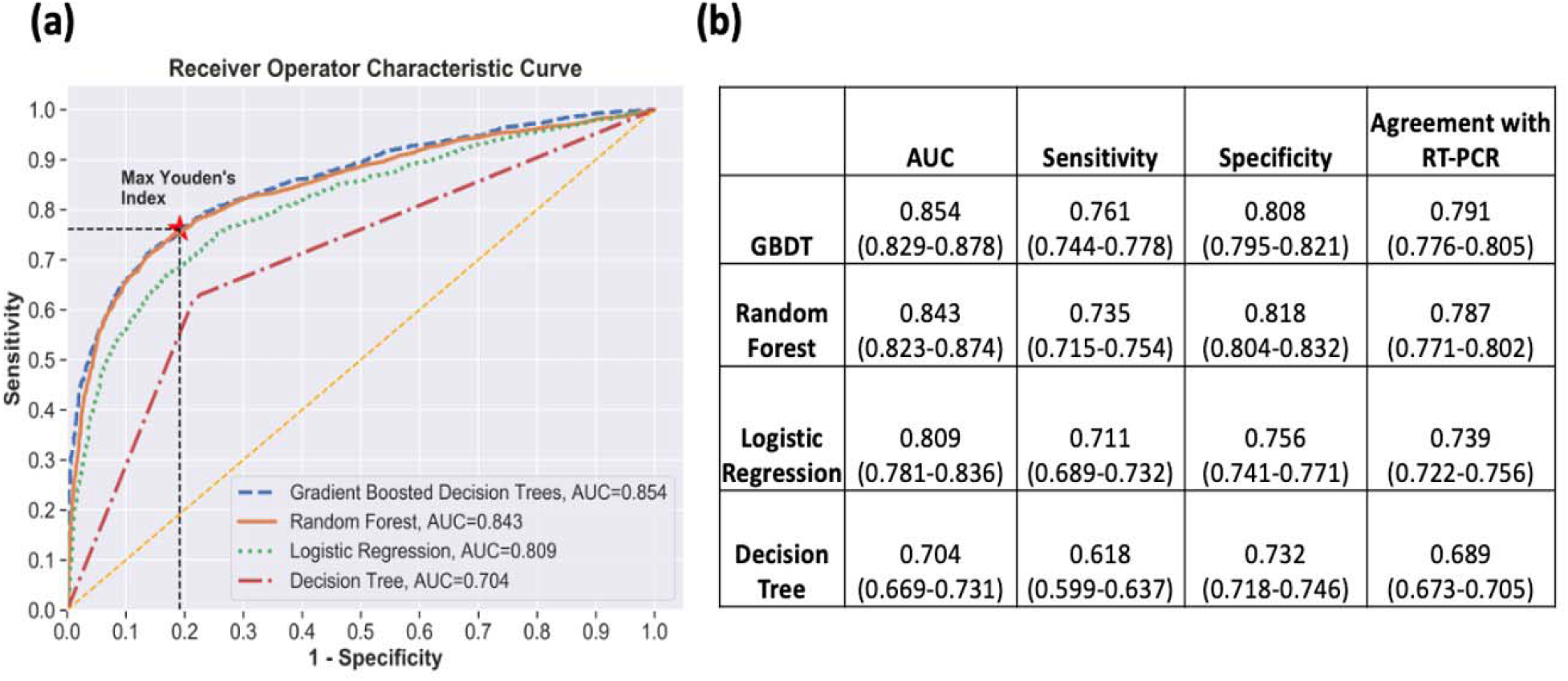
Performance of four models using five-fold cross validation on the test set. (a) Comparison of the ROC curves for the gradient boosted decision tree (GBDT) model, random tree model, logistic regression model, and decision tree model. (b) Comparison of the AUC, sensitivity, specificity, and agreement with SARS-CoV-2 RT-PCR (at the operating point determined by the Youden Index) achieved by the four models.

As an independent validation, we tested the performance of the trained model on an independent patient dataset (496 positive and 968 negative by RT-PCR, **Table 1**) collected from NYPH/LMH] during the same time period as the NYPH/WCM site. Using the model trained on NYPH/WCM patients, we were able to obtain an AUC of 0.838 on the NYPH/LMH data, confirming the ability to generalize our model to other hospitals. Given the same sensitivity (0.758) as NYPH/WCM, the specificity reached 0.740.

To further interpret the trained GBDT model, we adopted the Shapley additive explanations (SHAP) *(28)* technique using the SHAP package (https://github.com/slundberg/shap). It assigns each feature an importance value (the Shapley value) for each specific classification. The summary plot of the impact of laboratory tests to final prediction is shown in **Figure 4a**. For instance, higher lactic acid dehydrogenase (LDH) values drive a positive prediction, whereas lower lymphocyte count values drive a negative prediction. In addition, lower troponin values were seen in COVID-19 positive patients than the control group, who were also ill patients coming to hospital for other causes, such as myocardial infarction.

**Figure 4.**
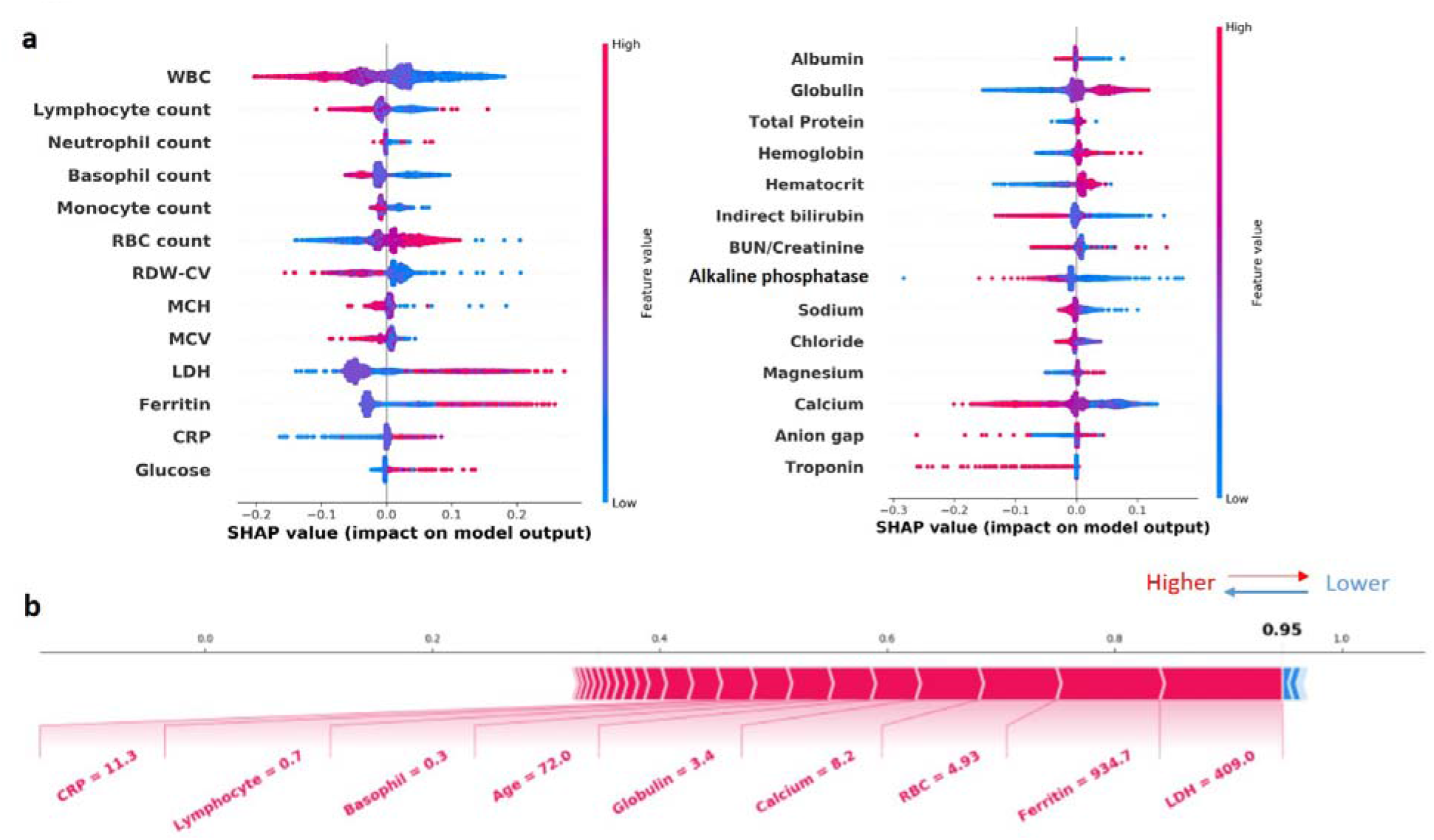
Contribution laboratory tests to the prediction of the GBDT model, using the SHAP technique. (a) The lab tests are organized on the y-axis according to their mean absolute SHAP values shown on the x-axis, which represents how the testing results drive the prediction of the GBDT model. Individual values of each test for each patient are colored according to their relative values. (b) Representative force plot of a patient who had a negative RT-PCR result at the initial ED visit. The GBDT model predicts a positive probability score of 0.95. Lab tests in red are positive “force” whereas test in blue are negative “force” driving the COVID-19 positive prediction.

Among the 32 patients whose SARS-CoV-2 RT-PCR results were initially negative but upon repeat testing within two days were positive, our approach predicted positive of the initial RT-PCR for 21 patients (66%). For example, a Hispanic male patient in his 70s underwent a RT-PCR testing which showed a negative result. Since the second RT-PCR test taken on the next day was positive, the initial RT-PCR was a suspicious false negative result possibly due to improper sample collection technique. The SHAP force plot (**Figure 4b**) illustrates how routine laboratory test results and demographics act as “forces” to push the GBDT model to make positive or negative predictions. The notable elevation of LDH, CRP, and ferritin in addition to lymphopenia and hypocalcemia in this patient, as well as his age, all contributed to a positive prediction by the GBDT model (positive probability = 0.95, 0.35 cutoff by the Youden Index), matching the subsequent RT-PCR result. Thus, our model may identify individuals with initial negative SARS-CoV-2 RT-PCR who should be retested and who could potentially need isolation at home or in the hospital while awaiting confirmatory RT-PCR results.

We also varied the length of the window for collecting routine lab tests with 4 more settings: one day before RT-PCR, one day after RT-PCR, one day before and one day after RT-PCR, two days before and one day after RT-PCR. The ROC curves are plotted in online **Supplemental Figure 2**. Analysis revealed that the longer the time window around the RT-PCR test, with more information captured characterizing patient infection status, resulted in slightly higher prediction performance in terms of AUC. However, these performances did not differ significantly from each other or from the chosen setting.

## Discussion

In this study, we proposed a machine learning model incorporating routine laboratory blood tests and a limited number of demographic features to predict an individual’s SARS-CoV-2 infection status. The model utilizes data that already exist in the medical record, rather than novel biomarkers, to predict SARS-CoV-2 infection status. Our model is able to generate a probability score of SARS-CoV-2 infection in real time before RT-PCR results are available, identifies the patients with a high risk of being SARS-CoV-2 positive and allows for quicker, more disease specific, patient management. As such, this model could be deployed clinically as an application integrated into the Electronic Medical Record (EMR) system. Using this application, clinicians could be alerted promptly of the infection risk level, allowing for rapid triaging and quarantining of high-risk patients as well as prompting rapid re-testing in those with positive model findings and negative SARS-CoV-2 RT-PCR results.

A recent study *(6)* proposed a machine learning model integrating chest CT findings with clinical symptoms, exposure history, leukocyte counts, age and sex to assist in the diagnosis of SARS-CoV-2 infection in a Chinese patient cohort. This model could be useful when CT scans are available for all patients. However, CT scans are not recommended as part of the initial routine clinical workup of COVID-19 by the American College of Emergency Physicians (ACEP) *(7)*. In contrast, our model is designed to complement the existing COVID-19 evaluation pathway based on the ACEP COVID-19 Field Guideline *(7)*. Predicting the probability of SARS-CoV-2 infection based on routine laboratory testing *without* radiology evidence is fast and inexpensive. However, it can be challenging because 1) the differences of individual laboratory test results are subtle between infected and non-infected patients at an early stage of the disease; and 2) in practice, patients may not undergo extensive laboratory testing, resulting in missing values in the dataset. In spite of these complications, our model has demonstrated a robust performance and delivers accurate predictions. More importantly, we have proven that the model can be generalized to other hospitals, other patient populations and other laboratory testing environments based on the predictability seen using an independent patient dataset. There have been attempts to build SARS-CoV-2 predictive models with routine laboratory tests *(10, 29)*. However, these studies were based on a small set of routine lab tests and the patient cohort sizes were smaller than our study.

The GBDT model was trained by iteratively selecting discriminative features from the root to leaf nodes and aggregating multiple trees with the weights determined from subset of the training samples. Thus, the tree nodes and the weights in the model reflect their impacts to the prediction, which is driven by the training data, making the model interpretable. For instance, larger values of inflammatory markers, such as LDH, ferritin, CRP drive to a positive prediction. These markers have been reported to be significantly associated with high risk of the development of COVID-19 and disease progression *(9, 30)*. Smaller values of lymphocyte count drive to a negative prediction. Lymphopenia is observed in a proportion of COVID-19 patients at hospital admission *(7, 31)* and is common features in severe COVID-19 patients *(3, 32)*. It is noteworthy that the control group in the training set consisted of patients negative for COVID-19 but ill for other diseases. That may explain why smaller values of a few lab tests, such as troponin, ALK, indirect bilirubin, were seen in the SARS-CoV-2 positive group compared to the control groups.

There are three potential limitations to the use of this model. First, the model was trained on a dataset generated from a patient cohort who were in the hospital for moderate to life-threatening presentations of COVID-19. Thus, this model may not be applicable to mild COVID-19 cases. Second, the model was developed with a “control group” of ill patients in a metropolitan hospital for other causes. Thus, the model may need further refinement with different populations such as patients seen in a primary care office. Third, clinical application of the proposed model may require modification of laboratory testing practice to include tests that are not currently part of the institutional COVID-like illness (CLI) laboratory test panel.

Generally speaking, an ideal training set for a learning-based approach should cover the variability of samples across different demographic and geographic distributions, as well as comorbidities, facilities (e.g. ED, inpatients, out-patient clinics) and to follow their changes over time. In practice, any training set collected within a fixed time period cannot satisfy all these wishes. The deployment of software in medical scenarios cannot be achieved by one stop. It is a continuous learning process that involves model monitoring, updating and customization. The US Food and Drug Administration (FDA) published a white paper *(33)* last year particularly discussing how to properly regulate the adaptations/modifications of AI/machine learning models as a medical device. The proposed ensemble model predicts COVID-19 infection by aggregating the decisions of a set of individual base learners, which are decision trees in GBDT. Because of such an additive nature, our ensemble model framework not only provides a convenient way of incorporating necessary model updating, but also makes model monitoring, maintenance, and regulation more flexible. New base learners should be trained on the new data when the model performance drops below a certain threshold due to the changes of patient population and testing strategies, and the ensemble model should be updated accordingly by incorporating the new base learner. The hyperparameters in the model, such as model updating frequency, model performance cutoff, number of base learners, etc., need to be determined according to concrete deployment scenarios.

## Conclusions

The proposed machine learning model, using age, gender, race and 27 routine laboratory tests, is a feasible and promising technique that can provide a rapid and objective prediction of SARS-CoV-2 infection status. The robust performance of our model was confirmed in an independent testing set. Our results have illustrated the potential role for this model as a tool to preliminarily identify high-risk SARS-CoV-2 infected patients before their RT-PCR results are available, risk stratify patients in the ED, and select patients who need relatively urgent re-testing, if the initial RT-PCR result is negative. Such use of our model could result in earlier appropriate isolation, thereby promoting the health of the patients while protecting the health of the public. Furthermore, our model may play an important role in assisting in the identification of SARS-COV-2 infected patients in areas where RT-PCR testing is not possible due to financial or supply constraints.

## Data Availability

The patient data used in this study were collected from NYP-WCM and NYP-LMH and thus not publicly available

## Acknowledgement

We want to thank Richard Fedeli for organizing the datasets of laboratory testing results.

## Author contribution

HSY for conceptualization, investigation, data analysis, writing, reviewing and editing of the manuscript, and visualization. YH for performing data analysis, investigation, reviewing and editing of the manuscript; LVV for organizing NYPH/LMH data; PS for editing the manuscript and providing ED information; AC for editing the manuscript; PV for providing RT-PCR information and performing analysis; SER, MMC, ML, RK for editing the manuscript; ZZ for conceptualization and editing the manuscript; FW for conceptualization, investigation, data analysis, visualization, editing the manuscript, and supervision of project.

## Funding/Support

The work is partially supported by National Science Foundation under grant number 1750326 and 1716432, and Office of Naval Research under grant number N00014-18-1-2585

**Abbreviation**
COVID-19: corona virus disease-2019;
SARS-CoV-2: severe acute respiratory syndrome coronavirus 2;
TAT: turn-around time;
ED: emergency department;
ICU: intensive care unit;
RT-PCR: real-time reverse transcription polymerase chain reaction;
GBDT: gradient boosted decision tree;
HCP: healthcare personnel;
WBC: white blood cells;
RBC: red blood cells;
LDH: lactic acid dehydrogenase;
RDW-CV: Red blood cell distribution width;
ALT: Alanine aminotransferase;
AST: Aspartate aminotransferase;
ALK: Alkaline phosphatase;
BUN: Blood urea nitrogen;
MCH: Mean corpuscular hemoglobin;
MCV: Mean corpuscular volume;
aPTT: Activated partial thromboplastin time;
CRP: C-reactive protein;
INR: International normalized ratio;
PT: Prothrombin time;
AUC: Area under the receiver operating characteristic curve;
NYPH/WCM: New York Presbyterian Hospital/Weill Cornell Medicine;
NYPH/LMH: New York Presbyterian Hospital/Lower Manhattan Hospital.

---

**Supplemental Figure 1:**
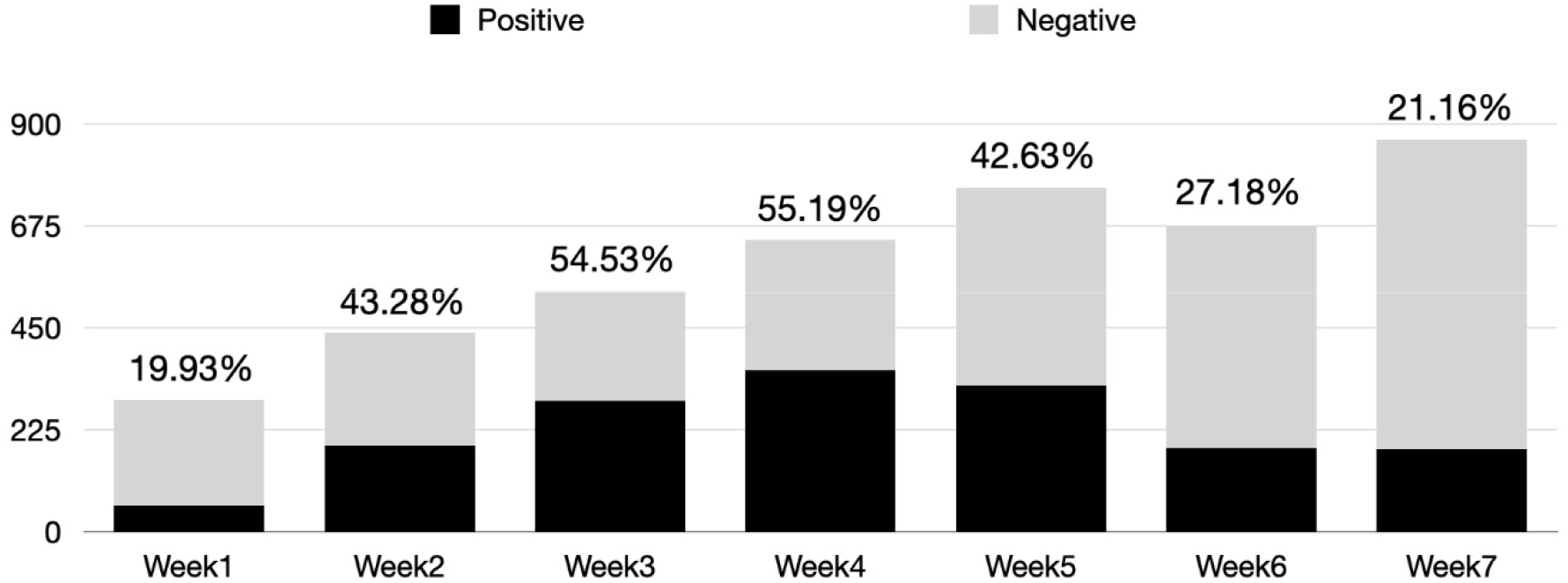
Bar chart of the number of RT-PCR tests performed weekly at NYPH/WCM from March 11 to April 29, 2020. Percent positive test rates are given at the top of each bar.

**Supplemental Figure 2.**
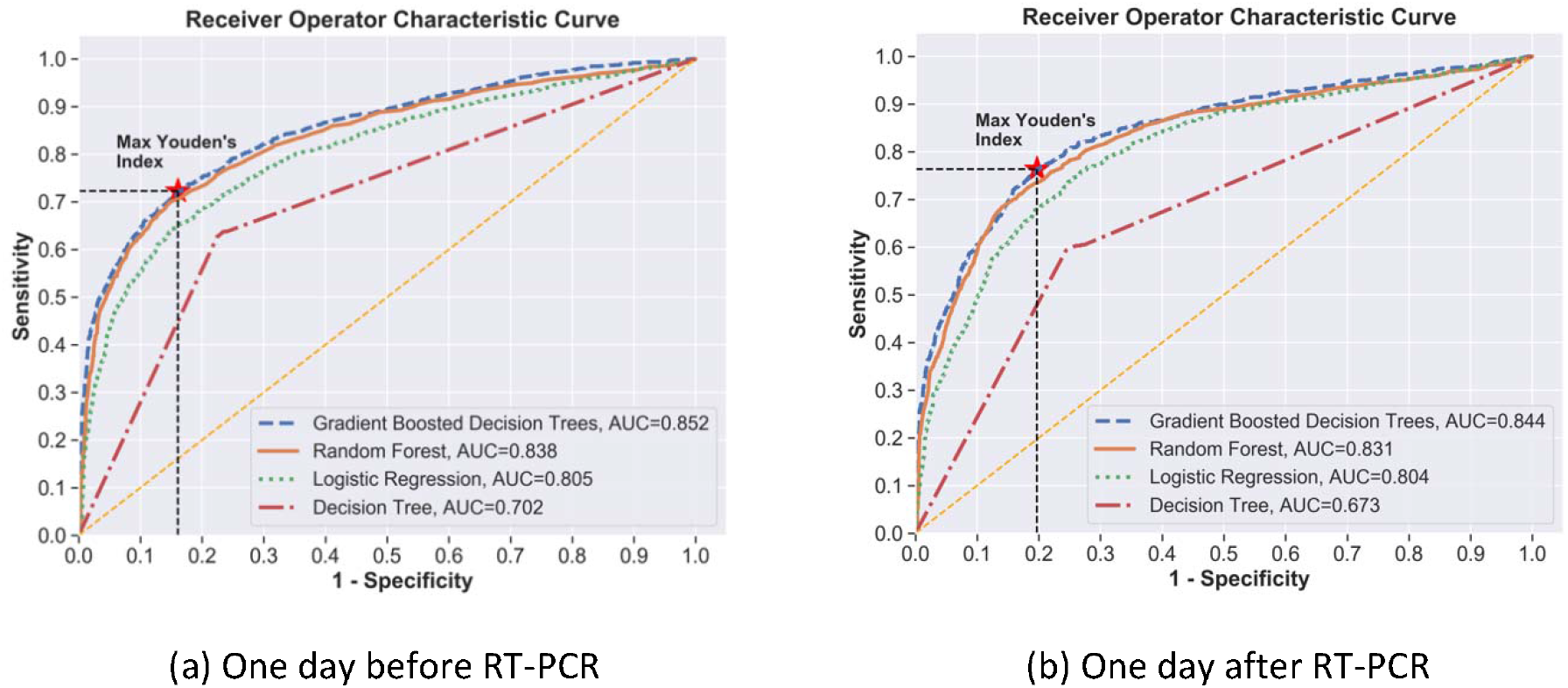
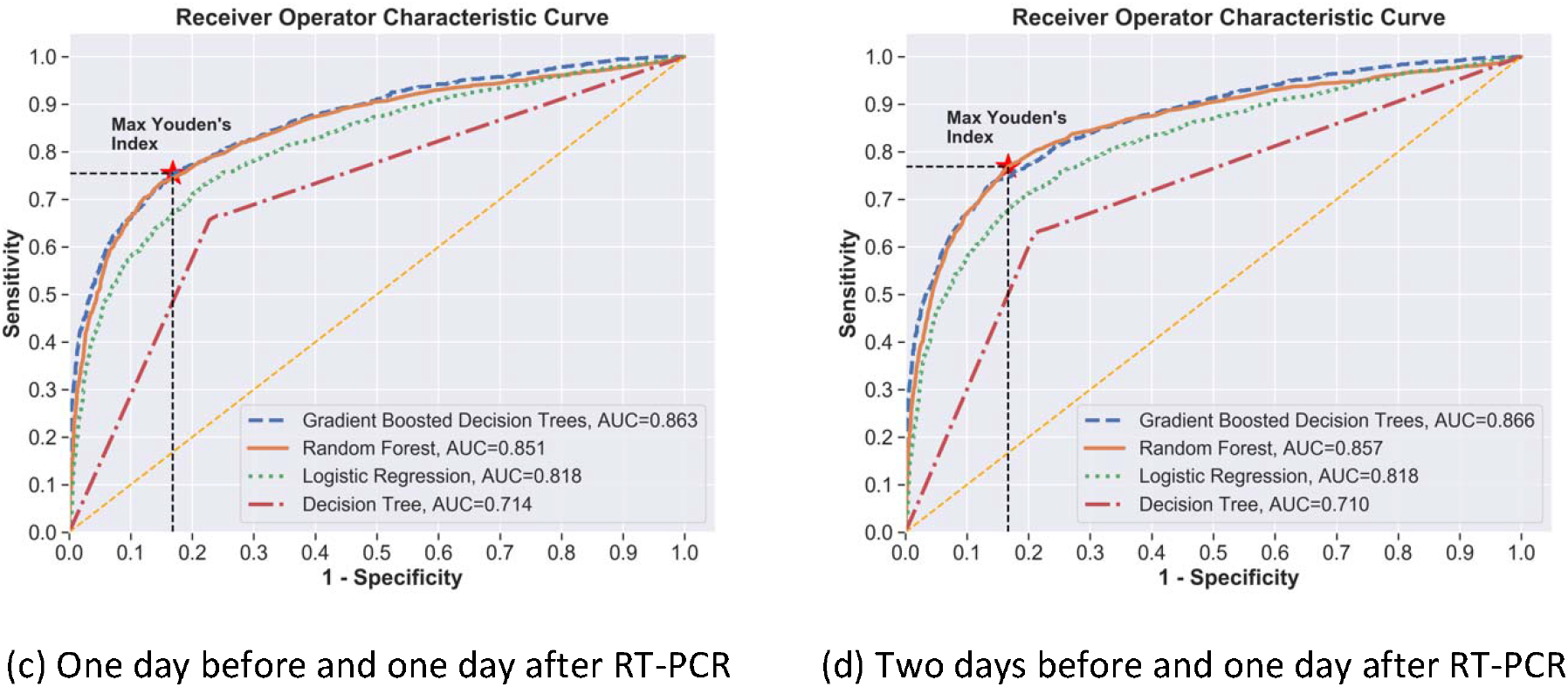
Performance of algorithms under different time windows in which routine lab test results were collected for the prediction of the RT-PCR result.

**Supplemental Figure 3.**
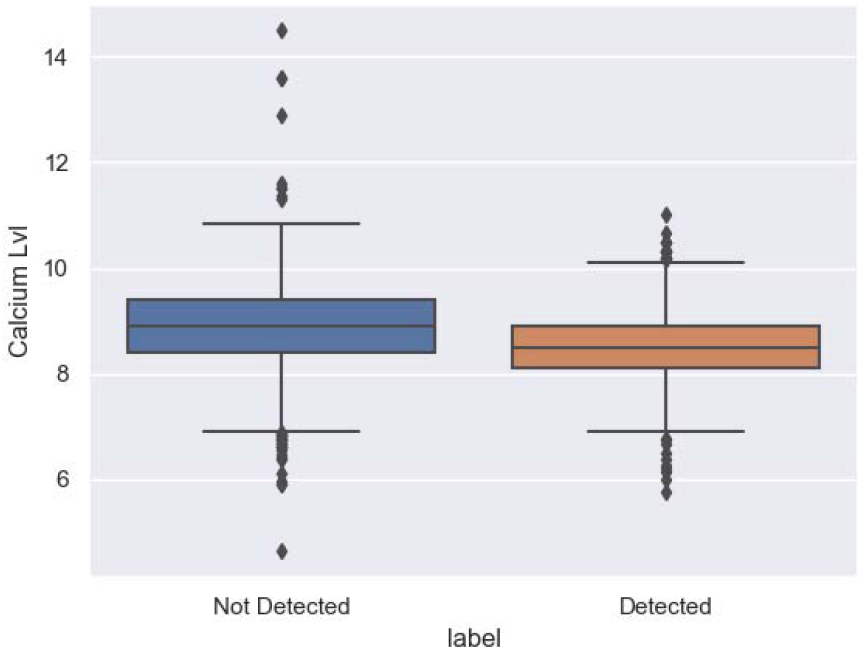
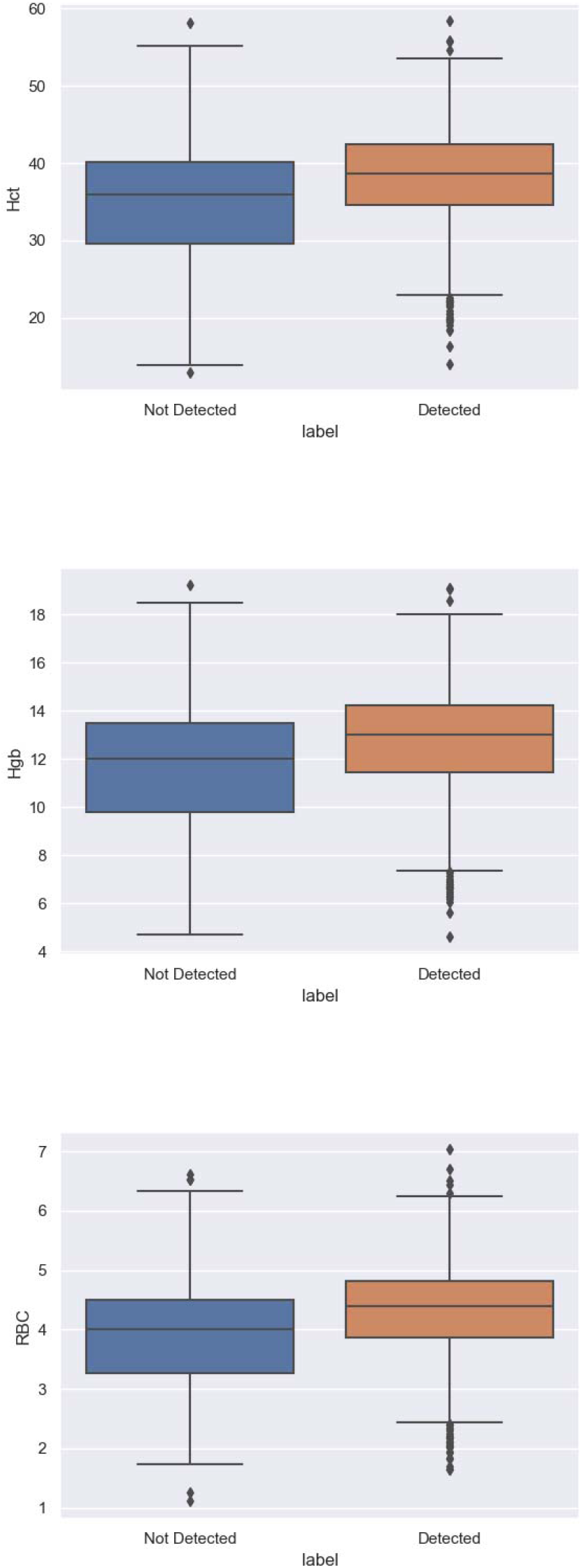
Representative box plots of laboratory tests.

**Supplemental Figure 4:**
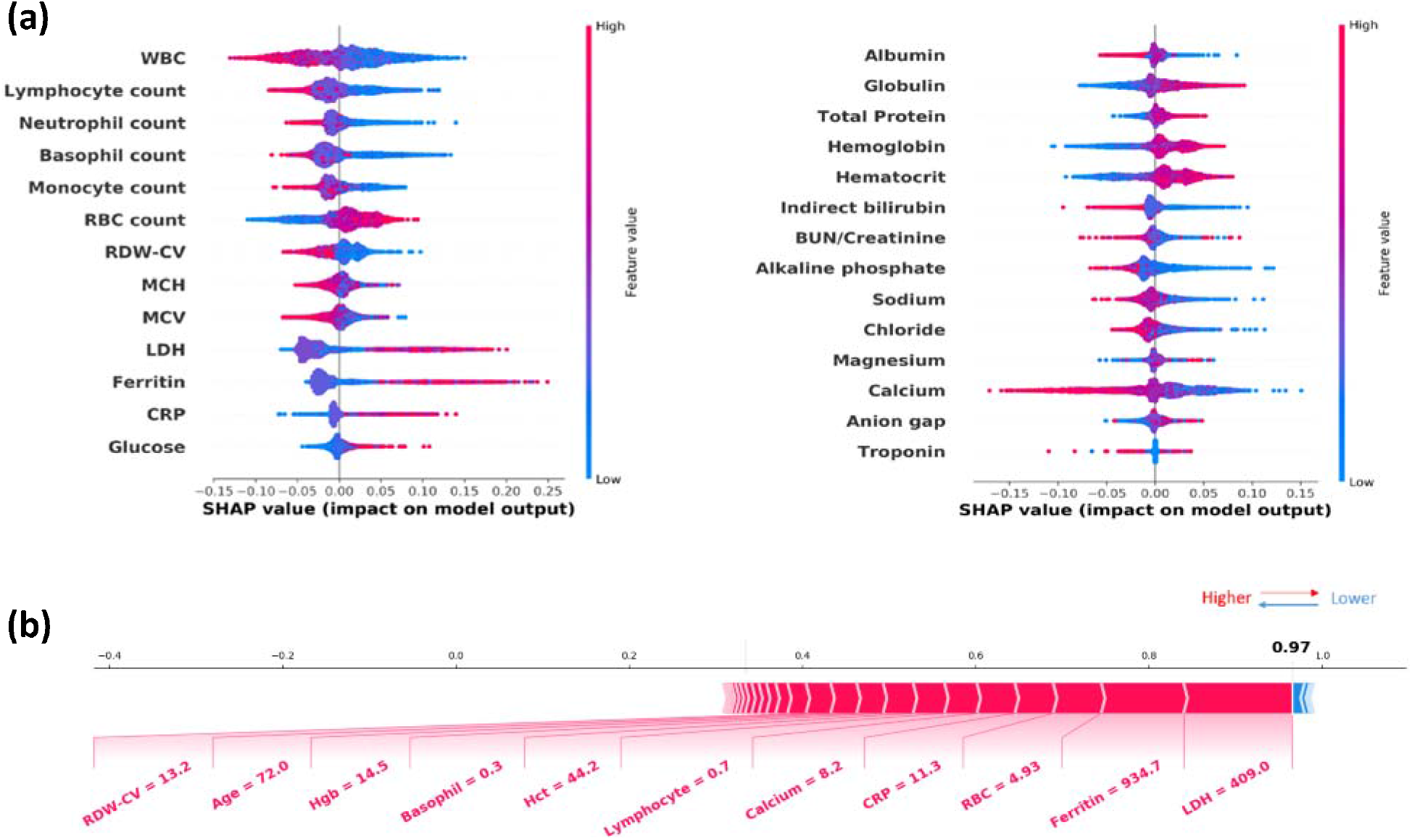
Variable importance measured by SHAP values as well as the explanation of the same case as shown in the paper using the Random Forest (RF) model. SHAP values of the routine lab tests obtained from the random forest (RF) model, which demonstrated similar patterns as they were obtained from GBDT (Figure 4a in the paper). (b) shows the force plot of the prediction made by the RF model on the same case demonstrated in Figure 4b in the paper, from which we can see RF predicts higher positive likelihood of this case (i.e., SARS-Cov-2 detected) with additional evidence from hemoglobin (Hgb), hematocrit (Hct) and red blood cell distribution width (RDW-CV).

**Supplemental Figure 5:**
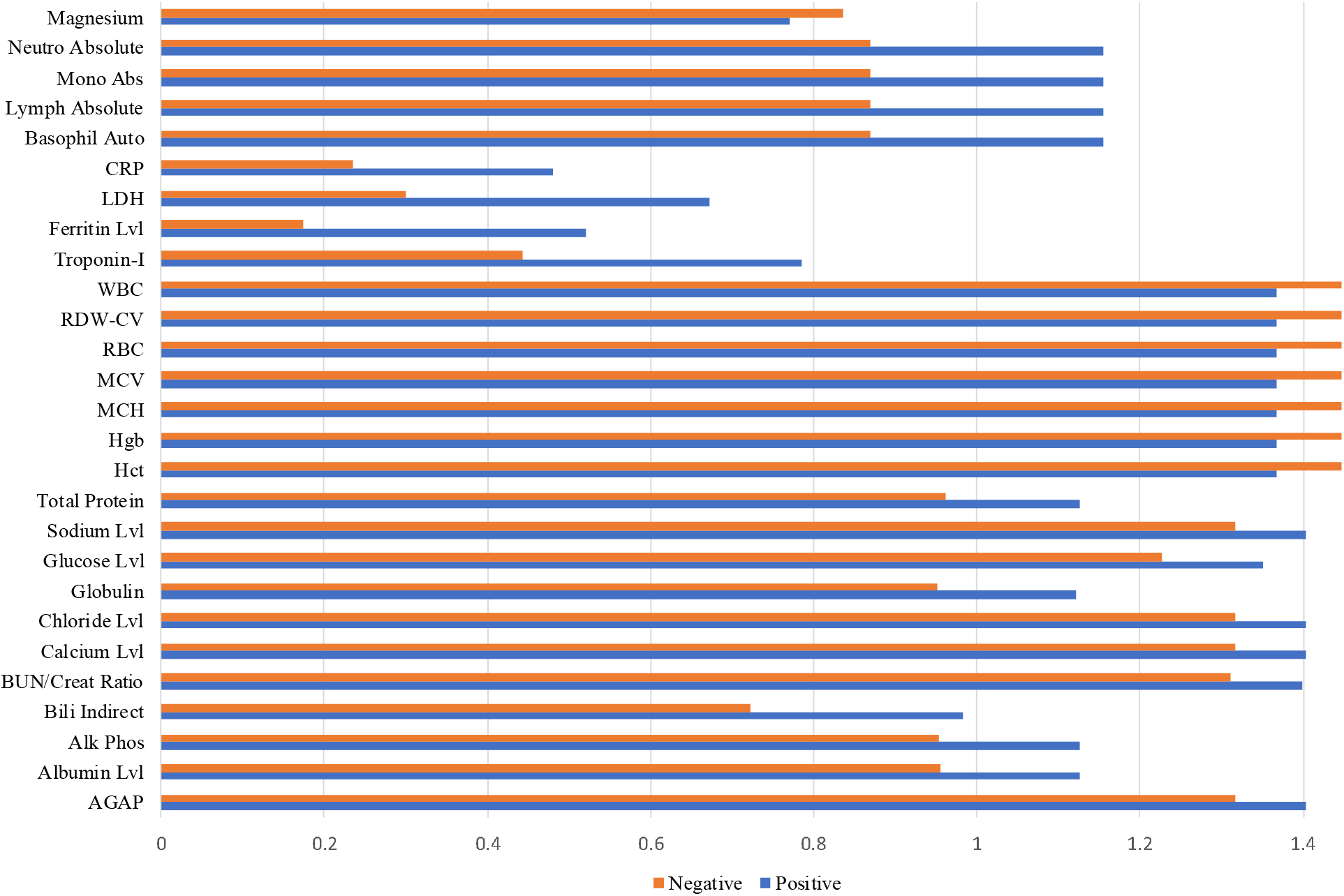
Comparison of testing frequency of each routine laboratory test within two days prior to each SARS-CoV-2 RT-PCR test (stratified by the positive and negative RT-PCR result)

**Supplemental Table 1:**
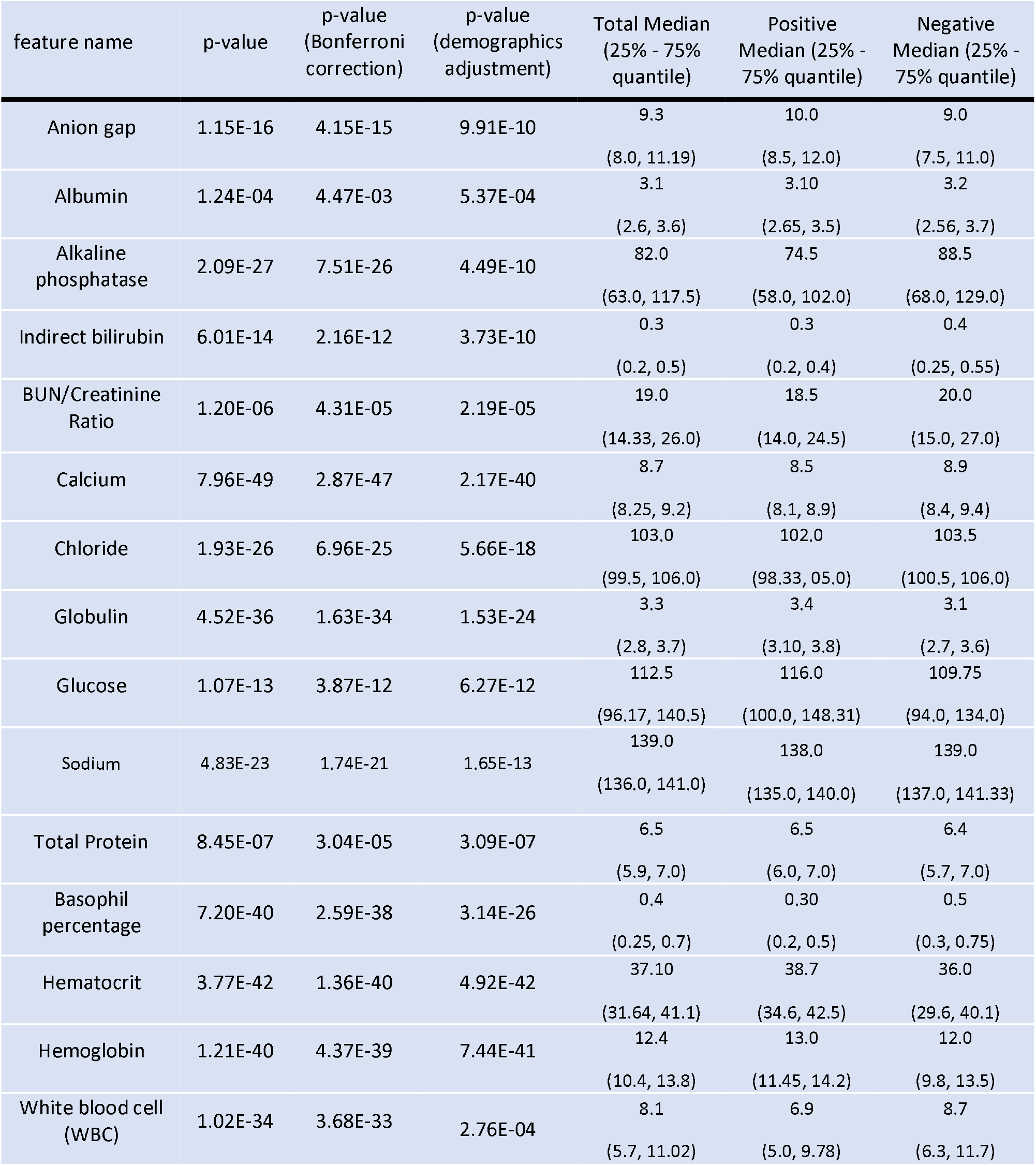
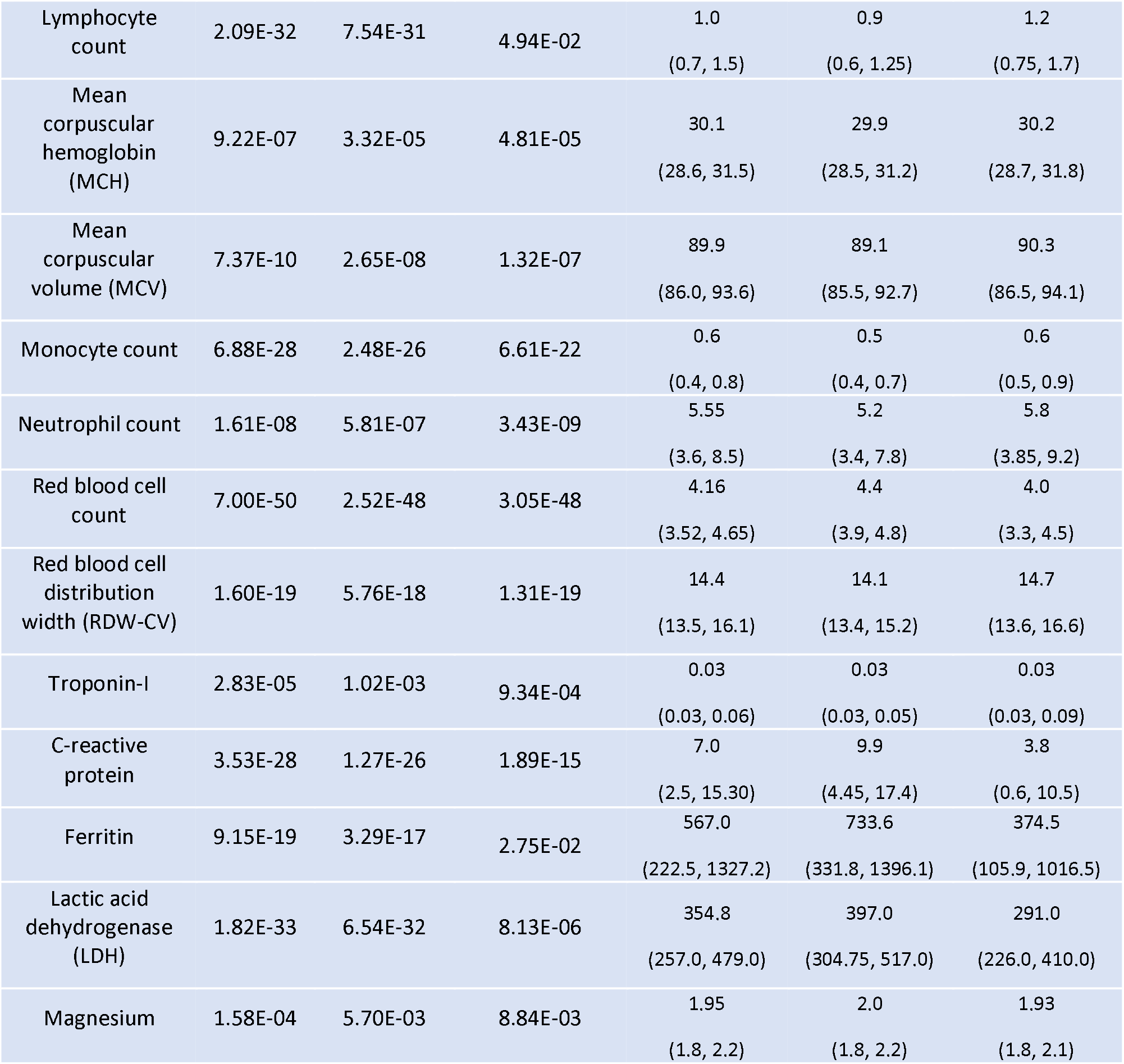
Summary of the statistics and P-values of 27 laboratory tests selected to construct the input feature vectors to the classification model. The demographic information used for *P*-value adjustment includes age, gender and race.

| feature name | p-value | p-value<br>(Bonferroni<br>correction) | p-value<br>(demographics<br>adjustment) | Total Median<br>(25% - 75%<br>quantile) | Positive<br>Median (25% -<br>75% quantile) | Negative<br>Median (25% -<br>75% quantile) |
| --- | --- | --- | --- | --- | --- | --- |
| Anion gap | 1.15E-16 | 4.15E-15 | 9.91E-10 | 9.3<br>(8.0, 11.19) | 10.0<br>(8.5, 12.0) | 9.0<br>(7.5, 11.0) |
| Albumin | 1.24E-04 | 4.47E-03 | 5.37E-04 | 3.1<br>(2.6, 3.6) | 3.10<br>(2.65, 3.5) | 3.2<br>(2.56, 3.7) |
| Alkaline<br>phosphatase | 2.09E-27 | 7.51E-26 | 4.49E-10 | 82.0<br>(63.0, 117.5) | 74.5<br>(58.0, 102.0) | 88.5<br>(68.0, 129.0) |
| Indirect bilirubin | 6.01E-14 | 2.16E-12 | 3.73E-10 | 0.3<br>(0.2, 0.5) | 0.3<br>(0.2, 0.4) | 0.4<br>(0.25, 0.55) |
| BUN/Creatinine<br>Ratio | 1.20E-06 | 4.31E-05 | 2.19E-05 | 19.0<br>(14.33, 26.0) | 18.5<br>(14.0, 24.5) | 20.0<br>(15.0, 27.0) |
| Calcium | 7.96E-49 | 2.87E-47 | 2.17E-40 | 8.7<br>(8.25, 9.2) | 8.5<br>(8.1, 8.9) | 8.9<br>(8.4, 9.4) |
| Chloride | 1.93E-26 | 6.96E-25 | 5.66E-18 | 103.0<br>(99.5, 106.0) | 102.0<br>(98.33, 05.0) | 103.5<br>(100.5, 106.0) |
| Globulin | 4.52E-36 | 1.63E-34 | 1.53E-24 | 3.3<br>(2.8, 3.7) | 3.4<br>(3.10, 3.8) | 3.1<br>(2.7, 3.6) |
| Glucose | 1.07E-13 | 3.87E-12 | 6.27E-12 | 112.5<br>(96.17, 140.5) | 116.0<br>(100.0, 148.31) | 109.75<br>(94.0, 134.0) |
| Sodium | 4.83E-23 | 1.74E-21 | 1.65E-13 | 139.0<br>(136.0, 141.0) | 138.0<br>(135.0, 140.0) | 139.0<br>(137.0, 141.33) |
| Total Protein | 8.45E-07 | 3.04E-05 | 3.09E-07 | 6.5<br>(5.9, 7.0) | 6.5<br>(6.0, 7.0) | 6.4<br>(5.7, 7.0) |
| Basophil<br>percentage | 7.20E-40 | 2.59E-38 | 3.14E-26 | 0.4<br>(0.25, 0.7) | 0.30<br>(0.2, 0.5) | 0.5<br>(0.3, 0.75) |
| Hematocrit | 3.77E-42 | 1.36E-40 | 4.92E-42 | 37.10<br>(31.64, 41.1) | 38.7<br>(34.6, 42.5) | 36.0<br>(29.6, 40.1) |
| Hemoglobin | 1.21E-40 | 4.37E-39 | 7.44E-41 | 12.4<br>(10.4, 13.8) | 13.0<br>(11.45, 14.2) | 12.0<br>(9.8, 13.5) |
| White blood cell<br>(WBC) | 1.02E-34 | 3.68E-33 | 2.76E-04 | 8.1<br>(5.7, 11.02) | 6.9<br>(5.0, 9.78) | 8.7<br>(6.3, 11.7) |
| Lymphocyte count | 2.09E-32 | 7.54E-31 | 4.94E-02 | 1.0<br>(0.7, 1.5) | 0.9<br>(0.6, 1.25) | 1.2<br>(0.75, 1.7) |
| Mean corpuscular hemoglobin (MCH) | 9.22E-07 | 3.32E-05 | 4.81E-05 | 30.1<br>(28.6, 31.5) | 29.9<br>(28.5, 31.2) | 30.2<br>(28.7, 31.8) |
| Mean corpuscular volume (MCV) | 7.37E-10 | 2.65E-08 | 1.32E-07 | 89.9<br>(86.0, 93.6) | 89.1<br>(85.5, 92.7) | 90.3<br>(86.5, 94.1) |
| Monocyte count | 6.88E-28 | 2.48E-26 | 6.61E-22 | 0.6<br>(0.4, 0.8) | 0.5<br>(0.4, 0.7) | 0.6<br>(0.5, 0.9) |
| Neutrophil count | 1.61E-08 | 5.81E-07 | 3.43E-09 | 5.55<br>(3.6, 8.5) | 5.2<br>(3.4, 7.8) | 5.8<br>(3.85, 9.2) |
| Red blood cell count | 7.00E-50 | 2.52E-48 | 3.05E-48 | 4.16<br>(3.52, 4.65) | 4.4<br>(3.9, 4.8) | 4.0<br>(3.3, 4.5) |
| Red blood cell distribution width (RDW-CV) | 1.60E-19 | 5.76E-18 | 1.31E-19 | 14.4<br>(13.5, 16.1) | 14.1<br>(13.4, 15.2) | 14.7<br>(13.6, 16.6) |
| Troponin-I | 2.83E-05 | 1.02E-03 | 9.34E-04 | 0.03<br>(0.03, 0.06) | 0.03<br>(0.03, 0.05) | 0.03<br>(0.03, 0.09) |
| C-reactive protein | 3.53E-28 | 1.27E-26 | 1.89E-15 | 7.0<br>(2.5, 15.30) | 9.9<br>(4.45, 17.4) | 3.8<br>(0.6, 10.5) |
| Ferritin | 9.15E-19 | 3.29E-17 | 2.75E-02 | 567.0<br>(222.5, 1327.2) | 733.6<br>(331.8, 1396.1) | 374.5<br>(105.9, 1016.5) |
| Lactic acid dehydrogenase (LDH) | 1.82E-33 | 6.54E-32 | 8.13E-06 | 354.8<br>(257.0, 479.0) | 397.0<br>(304.75, 517.0) | 291.0<br>(226.0, 410.0) |
| Magnesium | 1.58E-04 | 5.70E-03 | 8.84E-03 | 1.95<br>(1.8, 2.2) | 2.0<br>(1.8, 2.2) | 1.93<br>(1.8, 2.1) |

**Supplemental Table 2:** Model performance with different methods for aggregating the routine laboratory test values.

| Methods | AUC | Sensitivity | Specificity | Agreement with<br>RT-PCR |
| --- | --- | --- | --- | --- |
| mean | 0.8503<br>(0.8223, 0.8732) | 0.7526<br>(0.7363, 0.7683) | 0.8084<br>(0.7949, 0.8212) | 0.7874<br>(0.7728-0.8013) |
| median | 0.8494<br>(0.8223, 0.8729) | 0.7518<br>(0.7332, 0.7696) | 0.8079<br>(0.7946, 0.8207) | 0.7868<br>(0.7715, 0.8015) |
| min | 0.8493<br>(0.8218, 0.8725) | 0.7546<br>(0.7367, 0.7718) | 0.8029<br>(0.7889, 0.8163) | 0.7847<br>(0.7692, 0.7995) |
| max | 0.8510<br>(0.8228, 0.8729) | 0.7565<br>(0.7382, 0.7741) | 0.8043<br>(0.7907, 0.8174) | 0.7863<br>(0.7709, 0.8011) |
| mean_<br>change | 0.8528<br>(0.8228, 0.8758) | 0.7565<br>(0.7387, 0.7737) | 0.8086<br>(0.7947, 0.8219) | 0.789<br>(0.7736-0.8038) |
| mean_<br>frequency | 0.8515<br>(0.8244, 0.8725) | 0.7538<br>(0.7351, 0.7717) | 0.8098<br>(0.7961, 0.8228) | 0.7887<br>(0.7731-0.8036) |

**Supplemental Table 3:** Model performance with different imputation methods. **Feature Selection Process**. Univariate analysis was performed on all laboratory test results to obtain the significance of the association between each laboratory test and the RT-PCR result with SciPy1.4.1 (https://scipy.org/). Laboratory tests were selected to construct the input feature vectors of the prediction model based on the following criteria: 1) a result available for at least 30% of the patients two days before a specific SARS-CoV-2 RT-PCR test, and 2) showing a significant difference (*P*-value, *P*-value after Bonferroni correction, *P*-value after demographics adjustment all less than 0.05) between patients with positive and negative RT-PCR results. Supplemental Table 1 shows the details of the results. For laboratory tests with continuous values, we applied the Mann-Whitney U test using the scipy.stats.mannwhitneyu function in SciPy1.4.1. For laboratory tests with nominal values, we applied the chi-square test using scipy.stats.chisquare in SciPy 1.4.1. The Fisher’s exact test was performed if the number was too small (<5) for certain values of the laboratory test using the fisher.test function in R. For laboratory tests with continuous values, we applied the Mann-Whitney U test using the scipy.stats.mannwhitneyu function in SciPy1.4.1. For laboratory tests with nominal values, we applied the chi-square test using scipy.stats.chisquare in SciPy 1.4.1. The Fisher exact test was performed if the number was too small (<5) for certain values of the laboratory test using the fisher.test function in R. Laboratory tests having redundant clinical information were eliminated. For example, for neutrophil count and percentage, only the absolute neutrophil count was retained for analysis. After this procedure, 27 routine laboratory tests were selected for further analysis.

| Method | AUC | Sensitivity | Specificity | Agreement with RT-PCR |
| --- | --- | --- | --- | --- |
| median | 0.8503<br>(0.8240, 0.8727) | 0.7526<br>(0.7339, 0.7705) | 0.8084<br>(0.7949, 0.8213) | 0.7874<br>(0.7719, 0.8022) |
| mean | 0.8506<br>(0.8231, 0.8738) | 0.7552<br>(0.7367, 0.7730) | 0.8053<br>(0.7916, 0.8183) | 0.7864<br>(0.7709-0.8012) |
| KNN | 0.8251<br>(0.7943, 0.8471) | 0.7248<br>(0.7045, 0.7444) | 0.7833<br>(0.7695, 0.7967) | 0.7613<br>(0.745, 0.777) |
| random | 0.8034<br>(0.7100, 0.8370) | 0.7110<br>(0.6907, 0.7307) | 0.7477<br>(0.7339, 0.7612) | 0.7339<br>(0.7176, 0.7497) |

